# Sources of self-efficacy for physical activity among young adults in the United Kingdom: a mixed-methods investigation

**DOI:** 10.1101/2025.06.07.25329197

**Authors:** Phillip M. Gray, Andrew L. Evans, Hannah Bielby, Jamie C. Gillman

## Abstract

This study aimed to explore the relationships between sources of self-efficacy and self-efficacy for physical activity (PA), and compare interactions between mastery experience and other sources, among young adults in the United Kingdom. It also aimed to compare perceptions of sources among individuals of different mastery experience. A mixed-methods approach was employed. An analytical cross-sectional design was utilised, with 249 participants completing a questionnaire assessing sources and self-efficacy for PA. A subsample of 70 participants completed a qualitative questionnaire. Hierarchical linear regression and a coding reliability thematic analysis was conducted to analyse the quantitative and qualitative data, respectively. Mastery experience (*B* = 61.79, t(235) = 6.27, p < .001), vicarious experience (*B* = 20.75, t(235) = 2.56, p = .011), and self-persuasion (*B* = 43.90, t(235) = 6.29, p < .001), were significantly associated with self-efficacy for PA. The interactions between mastery experience and vicarious experience (*B* = -6.25, t(235) = -2.59, p = .010) and mastery experience and self-persuasion (*B* = -11.77, *t*(235) = -5.38, *p* < .001) were significant, indicating that the positive effects of vicarious experiences and self-persuasion on self-efficacy for PA weakened as mastery experiences increased. Other theorised sources were not associated with self-efficacy for PA. Qualitative findings generally supported and elucidated these findings, and highlighted that vicarious experience may facilitate or debilitate self-efficacy for PA. This study identified several significant associations between self-efficacy and its sources, as well as their interdependencies. Implications for theory and practice are discussed.

## Introduction

Physical inactivity is a risk factor for early mortality and non-communicable diseases.^1^ In Western countries, physical inactivity is attributed to 9.3% of all-cause, 9.9% of cardiovascular, 10.5% of dementia, and 2.6-9.3% of cancer mortality.^2^ To reduce this disease burden, adults aged 19-64 years should engage in 150 minutes of moderate physical activity (PA) in bouts exceeding 10 minutes, or 75 minutes of vigorous per week.^1^ However, guideline attainment within the United Kingdom remains suboptimal at 66% among males and 61% among female adults.^3^ Thus, investigating the factors that enable PA participation is imperative.

Perceived self-efficacy plays a critical role in the initiation of new behaviours, including PA. Self-efficacy refers to the belief in one’s abilities to organise and execute the action required to achieve a particular goal or behaviour^4^ and is a central tenet of established theories in exercise psychology. Self-efficacy beliefs can vary according to their; level, which refers to the difficulty of the tasks (from simple to complex); strength, which indicates the degree of certainty one has in their abilities (from low to high confidence); and generality, which reflects the transferability of beliefs across contexts.^4^ Individuals with high self-efficacy tend to choose more challenging behaviours, expend greater effort, persist in the pursuit of goals, demonstrate more adaptive responses following the completion of tasks, and maintain behaviours when faced with challenges.^4^ These tendencies have been specifically observed in the PA domain.^5^ Thus, self-efficacy is consistently one of the strongest predictors of PA behaviour adoption^6^ and maintenance^7^ and has been shown to mediate the effects of PA behaviour change interventions.^8^

Self-Efficacy Theory^4^ suggests four main sources of self-efficacy. Mastery experience refers to a person’s previous experience of executing an action (successfully or unsuccessfully) and cognitive appraisal (e.g., internal or external attribution).^4^ Mastery experience is the most potent source of self-efficacy information, given that it is derived from authentic experience.^4^ Second, vicarious experience describes the social modelling process whereby observing similar others successfully executing a behaviour increases self-efficacy.^4^ Through a third source, verbal persuasion, individuals are assured by others that they possess the capability to perform an action. Verbal persuasion is theoretically a weak source of self-efficacy, and any self-efficacy derived from this source may be readily extinguished by disconfirming authentic experiences.^4^ Judgements of somatic and emotional states constitute the fourth source of self-efficacy, physiological and affective states. Prior to or during PA, individuals may be subject to positive or negative affect, aches or pains, and the interpretation of such states influences self-efficacy.^4^ Experimental, observational, and qualitative research in the PA domain provide empirical support for these theorised antecedents of self-efficacy. Evidence indicates that self-efficacy for PA is strengthened through successful execution of PA tasks,^9–12^ positive verbal persuasion,^9^ vicarious experiences,^13^ and optimal physiological and affect states.^11,14^ Finally, research highlights self-persuasion (i.e. self-talk), as an important fifth source of self-efficacy.^10,15^ Moreover, the efficacy derived from these sources may depend on mastery experience. For instance, vicarious experience may be particularly important during the initial stages of behaviour adoption by providing a reference point^4^ whereas verbal persuasion may augment self-efficacy among those with established mastery experience.^12,16^

A scarcity of research has investigated the relative importance of self-efficacy sources,^16,17^ and ascertaining the relative predictive power of each may inform interventions with parsimonious designs and potentially increase intervention effectiveness.^17^ Among older German adults, Warner et al.^11^ demonstrated that mastery experience, negative affective states, and self-persuasion predict self-efficacy for PA. Inconsistent with Self-Efficacy Theory, these three sources were similar in magnitude, and other theoretical sources (i.e. vicarious experience and verbal persuasion) did not emerge as significant predictors. However, given that the relative importance of self-efficacy sources may be moderated by the behaviour investigated and demographic factors,^17^ including age,^18^ their findings may not be generalisable to younger cohorts in Western countries. Moreover, a scarcity of research has explored the interaction effects between sources,^17^ and as noted previously, this is an important area of investigation given that efficacy derived from sources (e.g., vicarious experience) may depend on an individual’s mastery experience. To date, there is no indication of what sources best combine with mastery, or which sources should be promoted if mastery is not yet established.^17^ Finally, a dearth of mixed-methods research has specifically investigated self-efficacy sources in the PA domain. Drawing on qualitative methods to complement a quantitative approach may facilitate a comprehensive and deeper understanding of the relative importance of sources of self-efficacy within the PA domain.^19^ Mixed-method approaches integrate two sets of strengths (i.e. breadth and depth), while simultaneously compensating for the weaknesses of the other. The approach facilitates the elaboration, enhancement, and clarification of results from one method to another, as well as the assessments of convergence and corroboration by data triangulation.^19^

Therefore, the first aim of the present study was to identify sources of self-efficacy associated with, and their relative importance for self-efficacy for PA among younger adults. Guided by Self-Efficacy Theory and previous empirical findings, mastery experience was hypothesised to exhibit the strongest association with self-efficacy, followed sequentially by positive affective states, self-persuasion, vicarious experiences, and verbal persuasion (hypothesis 1). It was hypothesised that negative affective states would be negatively associated with self-efficacy for PA (hypothesis 2). The next aim was to explore the interactions between mastery experience and other sources. This was expected to account for a significant proportion of the variance in self-efficacy, with the positive association between verbal persuasion and self-efficacy being stronger for those with higher, compared to lower levels of mastery experience (hypothesis 3), with the inverse being true for other sources (e.g., vicarious experience; hypothesis 4). Finally, to triangulate and elucidate any interactions emerging from the quantitative findings, the study also aimed to qualitatively explore perceptions of sources and their comparative importance for self-efficacy for PA among individuals of different mastery experience. In sum, this is the first study within the PA domain to quantitatively explore the associations between self-efficacy and self-efficacy sources among younger adults, compare interaction effects, and triangulate and complement findings with qualitative methods, thereby providing a unique contribution to the literature.

## Methods

### Participants

Participants were recruited from Greater Manchester, England, and self-selected through a combination of convenience and snowball sampling approaches. This was achieved by placing adverts in, or utilising gatekeepers of community groups, a university, social media, and word of mouth. Individuals were eligible for the study if they; a) were aged 18-64 years old; b) could understand and write in English, and c) provided written consent. Taking cognisance of the quantity of predictor variables in the current study, a minimum sample size of 154 (50+8 * number of predictor variables) was aimed to be recruited.^20^

Two-hundred and forty-nine individuals volunteered for the study. The sample had a mean age of 22.38 (± 6.54) years and was predominately male (60.2%) and educated to an undergraduate degree level (55%). Seventy-six percent of participants were White British or Irish, 10% were Asian or Asian British, 10% Black, African, Caribbean, or Black British, and 4% mixed ethnicity. A subsample of 70 participants were selected via simple random sampling to participate in the qualitative inquiry, which exceeds a minimum sample size likely to achieve data saturation.^21^ The subsample had a mean age of 22.73 (± 2.43) years and were predominantly male (63%) and educated to an undergraduate degree level (57%). Seventy-four percent of the subsample were White British or Irish, 11% Asian or Asian British, 10% Black, African, Caribbean, or Black British, and 5% mixed ethnicity.

Prior to data collection, ethical approval was obtained from the Ethics Committee of the School of X, University of X (approval number: HST1920-202) and informed consent was provided by participants.

### Protocol

#### Design

A pragmatic approach was adopted. Pragmatism does not commit to a particular philosophical stance, rather it values methodological pluralism and flexibility guided by the research aims. It values both objective and subjective knowledge, and the practical application of findings.^23^ A convergent mixed-methods design was employed. First, an analytical cross-sectional design was used to identify the associations between self-efficacy and sources of self-efficacy, and the interaction between sources. Second, a qualitative descriptive approach was used whereby information from those experiencing the phenomenon under investigation was directly elicited.^24^

#### Procedure

The participants attended the University of X for data collection once. Data collection was undertaken in a quiet and comfortable room, with a researcher present (P. G. or H. B.) who had no prior relationship with participants. P. G. possesses 13 years of experience in mixed-methods inquiries within research and practice and holds a PhD in exercise psychology. H.B. is an undergraduate student in Nutrition and Exercise as Medicine. The session began with introductions and an overview of the research and ethical considerations, including confidentiality and the right to withdraw. Next, participants completed a paper and pencil quantitative questionnaire. Following completion, participants were invited to undertake a voluntary review of the questionnaire to check for missing responses to items. If required, a clarification of these items was provided by the researcher. Participants selected for the qualitative subsample additionally completed an appended qualitative questionnaire. Data collection typically took between 30 and 45 minutes.

#### Measures

A questionnaire was used to collect sociodemographic information including age, sex, educational attainment, and ethnicity.

Self-efficacy sources were assessed using six subscales of the Sources of Self-Efficacy for Physical Activity Scale.^11^ The subscales measured a distinct source of self-efficacy: mastery experience, vicarious experience, verbal persuasion, positive affective states, negative affective states, and self-persuasion. Each item was answered on a 4-point Likert scale ranging from 1 (strongly disagree) to 4 (strongly agree) and averaged to produce a score for that subscale ranging from 1 to 4. The psychometric properties of the questionnaire have been established previously.^11^ As depicted in Table 2, the subscales had good internal consistency within the current study, with Cronbach’s alphas ranging from .75 to .82.

Self-efficacy for PA was assessed using the Exercise Self-Efficacy Scale (EXSE).^25^ The eight-item scale assessed participants’ beliefs in their ability to engage in regular moderate-intensity exercise. Each item was rated on an 11-point percentage scale ranging from 0% (not at all confident) to 100% (highly confident), in 10% increments, with responses averaged to produce an overall score ranging from 0 to 100.

Systematic review evidence has identified the EXSE as one of the most used and theoretically grounded measures of self-efficacy for PA, with strong alignment with self-efficacy theory through the specification of exercise frequency, duration, and intensity.^26^ The psychometric properties of the scale have been established previously^25^ and demonstrated good internal consistency (Cronbach’s α = .97) in the current study.

Self-efficacy sources were qualitatively investigated using a structured questionnaire. Structured questionnaires with open-ended questions and response formats have been used extensively within the health domain, and when well-designed can elicit good quality data.^27^ The questionnaire, presented in Table 1, comprised five open-ended questions, each probing a specific source of self-efficacy. Each question (e.g., ‘In your view, regarding physical activity, does your mastery experiences [or lack of], influence your self-efficacy? Please explain your answer’) was followed by a text box, which enabled open-ended responses. Preceding these questions, the participants were provided with written definitions and example statements of self-efficacy and each source to aid their comprehension. Questions were pilot tested in a separate sample to assess comprehension, perceived clarity, and utility for data richness, and no refinements were required.

**Table 1.** Questionnaire items probing self-efficacy sources.

| Questions |  |
| --- | --- |
| 1 | In your view, regarding physical activity, does your **mastery experiences** (or lack of), influence your self-efficacy? Please explain your answer. |
| 2 | In your view, regarding physical activity, does **vicarious experiences** (or lack of), influence your self-efficacy? Please explain your answer. |
| 3 | In your view, regarding physical activity, does **verbal persuasion** (or lack of), influence your self-efficacy? Please explain your answer. |
| 4 | In your view, regarding physical activity, does **physiological and affective states** (or lack of) influence your self-efficacy? Please explain your answer. |
| 5 | In your view, regarding physical activity, does **self- persuasion** (or lack of), influence your self-efficacy? Please explain your answer. |

### Statistical Analysis

For the quantitative data analysis, firstly, descriptive data and correlation analyses were performed using SPSS 27. Subsequently, a three-step hierarchical linear regression was conducted, with self-efficacy as the dependent variable. In Step 1, sex and age were entered as predictor variables in the null model. Mastery experience, vicarious experience, verbal persuasion, self-persuasion, positive affective states, and negative affective states were added as predictor variables to the model in Step 2.

Interactions between mastery experience and other self-efficacy sources were entered as predictor variables in Step 3. The hierarchical regression analysis model comparisons and a model interpretation are based on an alpha of .05. Each step in the hierarchical regression was compared to the previous step utilising *F*-tests. The coefficients of the model in the final step were interpreted. Effect sizes for the hierarchical regression were interpreted using incremental Cohen’s f², calculated from the change in R² between successive models. Values of 0.02, 0.15, and 0.35 were interpreted as small, medium, and large effects, respectively. Prior to the analysis, several assumptions were checked. Normality was evaluated for each model using a Q-Q scatterplot, which compares the distribution of the residuals with that of a normal distribution. Homoscedasticity was evaluated for each model by plotting the model residuals against the predicted values^28^, with non-random distribution and curvatures indicating violation of this assumption.^20^ To identify influential points, studentised residuals were calculated, and the absolute values were plotted against the observation numbers with those greater than 3.12 in absolute value, the 0.999 quantile of a *t* distribution with 248 degrees of freedom, considered to have significant influence on the results of the model.^20^

For the qualitative analysis, a coding reliability thematic analysis was employed^29^, which emphasises the importance of objectivity and minimising researcher bias, realised through the use of codebooks, multiple independent coders, and quantification of coding reliability.^30^ Questionnaire responses were word processed and uploaded to NVIVO (version 1.6.1). Next, the approach of coding and calculating the agreement specified by O’Connor and Joffe^30^ was adopted. Intercoder agreement was undertaken to facilitate research team reflection and dialogue, improve coding standards, and enhance coding frame specification.^30^ Therefore, the inter-coder agreement process supported reflexive dialogue and helped ensure that theme development was not solely dependent on one researcher’s perspective. The process involved the research team developing the first draft of a coding frame, using both deductive (self-efficacy sources and data collection questions) and inductive means (data familiarisation), and subsequent discussions. A codebook with a list of codes, labels, definitions, and examples was then created. The first coder (P.G.), then applied these codes to all data. The data was segmented by P.G., who used their judgement to determine what constituted meaningful conceptual breaks.^30^ A cleaned file, with the codes removed was then shared with a second researcher (J.G.), who coded the data using the same segmented data. J.G. holds a PhD in Sport and Exercise Psychology and possesses 10 years of experience of conducting qualitative data analysis. Files were merged, and intercoder agreement was calculated for each code, with a minimum threshold of 0.8 deemed acceptable.^31^ All codes exceeded this threshold (mean = 0.89 ± 0.05), therefore no further codebook revisions were required. Finally, participants were categorised by tercile into low, medium, and high mastery groups. Subsequently, a matrix coding query was computed to compare themes among these subgroups.

## Results

### Quantitative Findings

There was no missing data for any questionnaire items. Descriptive statistics and intercorrelations for self-efficacy sources and self-efficacy for PA are presented in Table 2. Normal distribution, homoscedasticity, and influential points assumptions were satisfied.

**Table 2.** Descriptive Statistics and Correlations for Study Variables.

| Variable | *M* | *SD* | $\alpha$ | 1 | 2 | 3 | 4 | 5 | 6 | 7 | 8 |
| --- | --- | --- | --- | --- | --- | --- | --- | --- | --- | --- | --- |
| 1. Age | 22.38 | 6.54 |  | - |  |  |  |  |  |  |  |
| 2. Mastery experience | 3.05 | 0.68 | .80 | -.12 | - |  |  |  |  |  |  |
| 3. Vicarious experience | 2.88 | 0.57 | .75 | -.12 | .05 | - |  |  |  |  |  |
| 4. Verbal persuasion | 2.88 | 0.60 | .76 | -.11 | .19* | .33* | - |  |  |  |  |
| 5. Positive affective states | 2.87 | 0.49 | .81 | -.18* | .3* | .17* | .11 | - |  |  |  |
| 6. Negative affective states | 2.18 | 0.59 | .79 | -.01 | -.31* | -.00 | .03 | -.13* | - |  |  |
| 7. Self-persuasion | 2.89 | 0.55 | .82 | .07 | .21* | .03 | .15* | .07 | .05 | - |  |
| 8. Self-efficacy for physical activity | 83.61 | 18.49 | .97 | -.08 | .57* | .05 | .12 | .23* | -.26* |  | - |

Note: \* < 0.05

### Comparing Models

The *F*-test for Step 1 was not significant, *F* (2, 246) = 0.11, *p* = .86, Δ*R*^2^ = .00, indicating that sex and age did not account for a significant amount of variation in self-efficacy. The *F*-test for Step 2 was significant, *F* (6, 240) = 35.17, *p* < .001, Δ*R*^2^ = .47, indicating that adding mastery experience, vicarious experience, verbal persuasion, self-persuasion, positive affective states, and negative affective states explained an additional 47% of the variance in self-efficacy. The *F*-test for Step 3 was significant, *F* (5, 235) = 10.06, *p* < .001, Δ*R*^2^ = .09, suggesting that the interactions between mastery experience and other self-efficacy sources explained an additional 9% of the variation in self-efficacy. The addition of self-efficacy sources in Step 2 produced a large incremental effect, f² = 0.89. The addition of interaction terms in Step 3 produced a medium incremental effect, f² = 0.20.

### Model Interpretation

Mastery experience (*B* = 61.79, t(235) = 6.27, p < .001), vicarious experience (*B* = 20.75, t(235) = 2.56, p = .011), and self-persuasion (*B* = 43.90, t(235) = 6.29, p < .001), were significantly associated with self-efficacy for PA (Table 3). Finally, mastery experience x vicarious experience (*B* = -6.25, t(235) = -2.59, p = .010) and mastery experience X self-persuasion (*B* = -11.77, *t*(235) = -5.38, *p* < .001) interactions were significantly with self-efficacy for PA. Sex, age, verbal persuasion, positive affective states, negative affective states, and all other interactions did not significantly predict self-efficacy for PA (*p* values > 0.05).

**Table 3.** Hierarchical regression analysis for variables predicting self-efficacy for physical activity.

| Variable | *B* | *SE* | 95.00% CI | *t* | *p* |
| --- | --- | --- | --- | --- | --- |
| Step 1 |  |  |  |  |  |
| (Intercept) | 82.07 | 4.28 | [73.63, 90.50] | 19.16 | < .001 |
| Age | 0.08 | 0.18 | [-0.28, 0.43] | 0.43 | .667 |
| Sex | -0.49 | 2.40 | [-5.22, 4.25] | -0.20 | .839 |
| Step 2 |  |  |  |  |  |
| (Intercept) | 10.96 | 9.80 | [-8.34, 30.27] | 1.12 | .264 |
| Age | 0.15 | 0.14 | [-0.12, 0.42] | 1.06 | .288 |
| Sex | -0.23 | 1.84 | [-3.85, 3.38] | -0.13 | .899 |
| Mastery experience | 13.24 | 1.48 | [10.31, 16.16] | 8.92 | < .001 |
| Vicarious experience | 0.49 | 1.68 | [-2.82, 3.81] | 0.29 | .770 |
| Verbal persuasion | 0.05 | 1.64 | [-3.17, 3.28] | 0.03 | .974 |
| Negative affective states | -3.64 | 1.59 | [-6.76, -0.51] | -2.29 | .023 |
| Positive affective states | 3.72 | 1.96 | [-0.13, 7.57] | 1.90 | .058 |
| Self-persuasion | 8.56 | 1.69 | [5.23, 11.90] | 5.05 | < .001 |
| Step 3 |  |  |  |  |  |
| (Intercept) | -120.75 | 28.87 | [-177.62, -63.88] | -4.18 | < .001 |
| Age | 0.08 | 0.13 | [-0.17, 0.33] | 0.65 | .513 |
| Sex | -0.13 | 1.69 | [-3.46, 3.20] | -0.08 | .940 |
| Mastery experience | 61.79 | 9.86 | [42.36, 81.21] | 6.27 | < .001 |
| Vicarious experience | 20.75 | 8.11 | [4.76, 36.73] | 2.56 | .011 |
| Verbal persuasion | -7.46 | 6.18 | [-19.63, 4.72] | -1.21 | .229 |
| Negative affective states | -6.48 | 5.82 | [-17.95, 4.98] | -1.11 | .266 |
| Positive affective states | 4.50 | 7.04 | [-9.38, 18.37] | 0.64 | .524 |
| Self-persuasion | 43.90 | 6.98 | [30.15, 57.65] | 6.29 | < .001 |
| Mastery experience X vicarious experience | -6.25 | 2.41 | [-11.00, -1.50] | -2.59 | .010 |
| Mastery experience X verbal persuasion | 1.88 | 1.97 | [-1.99, 5.76] | 0.96 | .340 |
| Mastery experience X negative affective states | 0.81 | 1.86 | [-2.85, 4.47] | 0.43 | .664 |
| Mastery experience X positive affective states | -1.35 | 2.34 | [-5.96, 3.26] | -0.58 | .564 |
| Mastery experience X self-persuasion | -11.77 | 2.19 | [-16.08, -7.46] | -5.38 | < .001 |

### Qualitative Findings

The qualitative analysis yielded five main themes aligned with each efficacy source. To address a study aim, findings are presented across different mastery levels. Participant quotations are presented to illustrate the findings, with identifiable information replaced with pseudonyms.

### Mastery Experience

Sixty-four participants indicated that mastery experience was an important determinant of their self-efficacy. This source was viewed as important across mastery groups, as demonstrated in the following quotations:

> A lack of mastery experience can lower my self-efficacy in exercise because if I have not succeeded in workouts before, I may feel less confident in my ability to exercise effectively or stick to a routine (Timmy, low mastery).
>
> Successful experiences influences my self-efficacy because if I’m able to gain experience while succeeding it gives me a confidence and self-belief on being able to continually try new things (Oscar, medium mastery).
>
> Yes, mastery experiences are important, as once I begin to develop my knowledge and confidence in myself in physical activity, I will become even more motivated to commit and succeed (Niamh, high mastery).

For some participants, mastery experiences were important for building self-efficacy, however it did little to diminish other sources of self-efficacy when facing new tasks, as illustrated by the following excerpts:

> Yes, mastery experiences influences my self-efficacy. If I know I’ve been successful before when taking on a new challenge, it will help me believe that I can be successful again. However, I still feel a sense of nerves when stepping into the unknown (Jordan, medium mastery).

Physiological and Affective States: Fifty-seven participants indicated that both positive and negative physiological and affective states contributed to their self-efficacy. This was deemed an important source of self-efficacy, irrespective of mastery level. Notable examples include:

> Feeling energetic, healthy and positive can increase my confidence in exercising, while fatigue, stress or discomfort can lower it (Luke, low mastery).
>
> Yes, physiological and affective states influence my self-efficacy because in the right state of mind and body it increases my self-efficacy and believe that I can exercise successfully (Ciara, medium mastery).
>
> Physiological and affective states influences my self-efficacy definitely. If I’m feeling anxious or something my self-efficacy may be altered negatively (William, high mastery).

Self-Persuasion: Fifty-five participants stated that self-persuasion influenced self-efficacy, with this view endorsed universally among those with low mastery. Although most participants in the low mastery group described positive self-persuasion as enhancing self-efficacy, they were more likely than those with higher mastery to discuss the detrimental influence of negative self-persuasion. This is reflected in the following comments:

> Yes, I’m a bit negative about my ability therefore don’t always feel confident to try (Princess, low mastery).
>
> Self-persuasion influences my confidence, yes. If I tell myself, I can do something pushes me to work harder for it and increase my belief that I can do it (Holly, low mastery).

Among those with medium and high mastery levels, perceptions of the influence of self-persuasion were mixed, as exemplified in the following quotations:

> Self-persuasion does not influence my self-efficacy. I think I rely on outside factors to motivate and make myself believe I can complete the exercise (Thomas, medium mastery).
>
> No I don’t feel my self-efficacy is really altered by this (William, high mastery).
>
> Self-persuasion influences my self-efficacy, yes, because when I tell myself I can do something it gives me self-belief that I can do it well (Oscar, medium mastery).
>
> Definitely. I always use self-persuasion during exercise especially when it gets hard, to help me get through the session and give it 100%, whatever 100% looks like on the day (Natalia, high mastery).

### Vicarious Experience

Fifty-four participants indicated that vicarious experience influenced their self-efficacy. Congruent with Self-Efficacy Theory, some participants indicated that observing others with similar characteristics, augmented the efficacy derived from this source. This is articulated in the following comments:

> Yes, vicarious experiences influences my self-efficacy. If I see someone exercise and I feel they are similar capability or physical feats, then it makes me more confident that I can also do those physical feats (Muhammad, low mastery).
>
> Learning off others influences my self-efficacy. Watching people successfully completing a task gives me confidence in my ability to also complete that task successfully as I’ve been shown how to do it through observation (Edward, medium mastery).
>
> Observing others successfully complete a task has motivated me to try new activities, especially if they are a similar age. I do tend to think ‘well if they can do it so can I’, which has led to learning new skills such as learning front crawl at the age of 50 (Amy, high mastery).

However, this perceived positive influence was not universal. Several participants across mastery levels indicated that vicarious experiences undermined self-efficacy, as noted in the following excerpts:

> Yes, vicarious experiences influences my self-efficacy, but negatively. No matter how well other people perform, it never happens to me, as I never achieve greater than others when watching someone be too successful because when watching this, it demotivates me to think I can never achieve better (Noel, low mastery).
>
> Vicarious learning off others may influence my self-efficacy, either positively or negatively. Depending on the task, seeing someone else completing the task could make me nervous and undermine my confidence or perhaps sometimes more confident that I can perform the task (Liam, medium mastery).
>
> Yes, if I see someone succeeding at something that I cannot do, then my self-efficacy decreases and therefore do not want to try again, as I think I am a failure (Chloe, high mastery).

### Verbal Persuasion

Verbal persuasion was deemed a determinant of self-efficacy among forty-nine participants and was universal across mastery levels. These participants generally considered positive persuasion to enhance self-efficacy, and negative persuasion to undermine it. For example:

> Verbal persuasion influences my confidence as if someone is giving me positive feedback on physical activity performance it will enhance my belief in my ability, and vice versa if criticised on my performance, I will lack belief in my own ability when performing again (Holly, low mastery).
>
> Yes, very much. Receiving positive feedback is encouraging because it validates the effort I have put into something. It feels like my hard work is noticed and appreciated, which makes me want to work harder. Although negative feedback can be discouraging, it can also push me to improve because if others can do it, then so can (Callum, high mastery).

However, for other participants, verbal persuasion was less relevant, as illustrated by the following quotations:

> Not really, verbal persuasion from others does not influence my self-efficacy. I rarely get persuaded by others and most people in my social circle are inactive. Therefore, it’s me personally that has to persuade myself if I want to be active (Sonny, low mastery).
>
> No, I prefer independence most of the time. It can feel condescending, almost like I’m not truly capable (Sarah, high mastery).

## Discussion

To date, there has been a scarcity of research that has explored the associations between sources of self-efficacy and self-efficacy for PA among younger adults. Moreover, previous research has not examined the interactions between mastery experience and other sources of self-efficacy for PA, nor utilised qualitative methods to triangulate quantitative findings. Addressing these aforementioned gaps may yield important insights on how to increase the effectiveness and efficiency of PA interventions. Additionally, it may identify which sources of self-efficacy should be emphasised at varying levels of mastery. Using a mixed-methods approach, this study has satisfied these important research gaps.^11,16,17^ Mastery experience exhibited the strongest association with self-efficacy, followed sequentially by self-persuasion and vicarious experience. Verbal persuasion, positive affective states, and negative affective states were not significantly related to self-efficacy for PA. These findings partially support hypothesis 1, and do not support hypothesis 2. Moreover, an interaction was found between mastery experience and self-persuasion, and mastery experience and vicarious experience, with theses sources being more relevant to individuals with lower, compared to higher mastery experience. No interactions were found between mastery experience and any other sources of self-efficacy. These findings do not support hypothesis 3 and partially support hypothesis 4. Qualitative findings generally corroborated and provided insights into these findings.

The quantitative findings indicated that mastery experience has the strongest association with self-efficacy for PA for these participants. This was supported by the qualitative findings. Participants, irrespective of mastery level, reported mastery experience as important for developing PA-related knowledge and confidence, and enhancing commitment and willingness to attempt new challenging tasks. These findings are consistent with previous quantitative research among younger Western adults^12^ and also align with qualitative research among South Korean^32^ and Western^9^ middle-aged adults. Among older adults, mastery experience has also previously been shown to be significantly associated with self-efficacy for PA,^11^ albeit contrary to this study, was of similar magnitude to other sources. However, the current findings are consistent with the theoretical propositions of Self-Efficacy Theory, which contests mastery as the most potent source, given that efficacy information is derived from direct authentic experiences.^4^ Efficacy expectations are developed by integrating efficacy information derived from repeated successes, particularly when attributed to internal factors and ability, and when facing novel and optimally challenging tasks. In contrast, repeated failures undermine self-efficacy.

Self-persuasion was found to be significantly associated with self-efficacy for PA. This was corroborated by the qualitative findings, which highlighted its importance for building confidence, and maximising effort and motivation for PA tasks. Existing evidence has demonstrated self-persuasion as an important source of self-efficacy for PA.^15^ Its significance has been demonstrated even after accounting for other sources^11^ and may be more effectual if ‘motivational’ forms of self-persuasion are employed.^33^ Motivational self-persuasion, as measured in the current study, includes cues that aim to maximise effort, psyching up, building confidence, and emotional control (e.g., ‘I can do it’ and ‘keep going’.; ^34^], and is suggested to be more relevant for tasks that require endurance, strength, and gross motor skills, such as PA.^33^ Interestingly, the efficacy of self-persuasion was contingent on mastery experience. Specifically, the positive effects of self-persuasion on self-efficacy for PA weakened as mastery experiences increased. This finding was congruent with the qualitative data, which further suggested that self-persuasion may be especially influential among individuals with low mastery because it can operate in both adaptive and maladaptive directions. Prior research has demonstrated that self-persuasion is more beneficial for novel, rather than well-learned tasks,^34^ and may also be more useful for novice, as opposed skilled performers whose behaviours are more automatic and involve less cognitive activity.^33^

In contrast to self-persuasion, verbal persuasion was not found to be associated with self-efficacy for PA. With some notable exceptions,^9,10^ this finding aligns with other research in the PA domain, comprising of diverse designs and populations.^11–13^ Theoretically, verbal persuasion is a weak source of self-efficacy because it does not originate from authentic experiences. Self-efficacy derived from this source can be attenuated if an individual has prior or future adverse PA experiences.^4^ Participants’ perceptions of verbal persuasion were heterogeneous; some emphasised its importance, whereas others reported limited receptivity to, or lacked a social milieu from which PA- related persuasive messages could be derived.

These findings corroborate previous assertions that verbal persuasion may augment self-efficacy for certain individuals, but be ineffectual or detrimental to others.^35^ Negative or critical verbal persuasion in particular can lead to feelings of incompetence or discouragement, with the debilitating effects of persuasive information more powerful than the enabling effects.^4^ Moreover, verbal persuasion may be ineffective or detrimental when perceived as controlling, manipulative, or unauthentic.^11,16^

Vicarious experience was significantly related to self-efficacy, a finding consistent with certain research in the PA domain,^13,32^ but contrasting with others.^11,14^ The findings of the current study are consistent with Self-Efficacy Theory, which posits that observing others of similar characteristics successfully undertaking and persisting at a behaviour develops self-efficacy. The qualitative data suggests that while vicarious experience augments self-efficacy among certain individuals, for others, it engenders perceptions of inefficacy, failure, and demotivation. Theoretically, among individuals with weakened self-efficacy, this may result from observing the success of others with dissimilar characteristics (e.g., of enhanced skill set or baseline fitness levels) or arise from the mode which vicarious experience is derived. For instance, social media is used by over 90% of adults aged 16 to 44 years in the United Kingdom,^36^ and increasingly functions as a medium for PA-related social learning.^32^ However, PA-related social media content is often self-selected^32^ to present favourable or unrealistic outcomes, and omit critical information about the strategies, sustained effort, failures, difficulties, and coping strategies associated with attaining behaviour or behavioural outcomes, factors imperative for augmenting self-efficacy.^8^ An interaction was found between mastery experience and vicarious experience, whereby the positive effects of vicarious experiences on self-efficacy for PA weakened as mastery experience increased. This finding is consistent with Self-Efficacy Theory that suggests vicarious experience is a more pertinent source of self-efficacy among individuals where mastery has not yet been established.^4^

Finally, positive affective states and negative affective states were not quantitatively associated with self-efficacy. In contrast, participants of the current study emphasised the detrimental role of negative affective states, such as anxiety and stress in weakening self-efficacy, whereas positive affective states were reported to enhance it. Challenging tasks often evoke emotional arousal, and negative emotions, such as anxiety, often construed as indicators of unpreparedness or lacking the capability to perform a behaviour.^4^ Conversely, positive affective states can be interpreted as preparedness, which augments self-efficacy. Moreover, according to Bandura^4^, past successes and failures, along with their associated emotional states, are stored in the memory. Thus, negative moods can elicit recollections of past failures, whereas positive moods can activate memories of past accomplishments.

Theoretical and Practical Implications: The findings of the current study have theoretical implications. Certain findings were consistent with Self-Efficacy Theory. For example, mastery experience and vicarious experience were positively associated with self-efficacy, with the latter more pertinent for those with lower mastery. The findings however diverge from other propositions of the theory. For instance, verbal persuasion, positive affective states, and negative affective states were not significantly associated with self-efficacy for PA. The qualitative findings suggested that vicarious experience can be either facilitative or debilitative of self-efficacy. Finally, self-persuasion was positively related to self-efficacy for PA, and both the quantitative and qualitative findings suggest this is a source more pertinent to those of lower mastery. The large unexplained variance also suggests that additional sources beyond those proposed by Self-Efficacy Theory may be related to self-efficacy for PA.

Mastery experiences, self-persuasion, and vicarious experience were significantly associated with self-efficacy, and may warrant inclusion in future PA interventions. The findings suggest that self-persuasion and vicarious experience may be of greater utility for individuals with low, compared to established mastery. In other words, these efficacy sources may be more useful to increase self-efficacy when learning a new skill or exercise. This nuanced understanding of the self-efficacy sources is of particular importance to help aid PA participation and adherence via tailored interventions. These sources can be manipulated in interventions that aim to increase self-efficacy by employing aligned BCTs^37^, with numerous interacting BCTs likely to enhance effectiveness^35^. However, given the negative consequences of ‘overconfidence’, practitioners or researchers should refrain from intervening among those who already possess high self-efficacy^5,17^. First, mastery can be promoted by providing opportunities to practice PA behaviour (BCT 8.1: behavioural practice), with a progressive overload of frequency, intensity, duration, or modality (i.e. task complexity; BCT 8.7: graded tasks). Setting, monitoring, reviewing, and readjusting PA-related goals, either by oneself or an exercise professional, will help facilitate progressive overload (BCTs 1.1, 1.5, 2.1 and 2.3: goal setting behaviour, review of behavioural goal, monitoring of behaviour by others without feedback, and self-monitoring of behaviour). Moreover, scaffolded guidance, support, and feedback from an exercise professional (BCTs 2.2, 3.1, and 4.1: feedback on behaviour, social support, instruction on how to perform the behaviour), who additionally encourages a focus on past successful experiences of PA (BCT 15.3: focus on past success) and the reattribution of past or current failures (BCT 4.3: re-attribution) may also enhance mastery. Progressing these behaviours to a minimum frequency of four times per week^38^ within a stable context may facilitate habit formation (BCT 8.3: habit formation) or alternatively could be generalised to other contexts (BCT 8.6: generalisation of target behaviour).

To maximise self-efficacy derived from self-persuasion, individuals with low mastery should be encouraged to undertake motivational self-talk (BCT 15.4: self-talk) and provide relevant resources and training to do so. To enhance self-efficacy through vicarious experiences, interventions should incorporate demonstrations of the behaviour (BCT 6.1), and social comparison (BCT 6.2) whereby individuals observe similar others successfully performing and persisting with PA despite challenges.

Strengths and Limitations: Several limitations of the current study warrant consideration. Firstly, although both samples were generally representative of the United Kingdom’s population for ethnicity and educational attainment, both were disproportionally comprised of males, younger adults, and exclusively of individuals residing in the North-West of England.

Thus, readers should judge the transferability of findings to other populations, or consider replicating the study using probability sampling methods to ensure a more representative sample. As noted by Bandura^4^, the ability to discern, weight, and integrate efficacy information improves with the development of cognitive and meta-cognitive skills. Therefore, the findings may not reflect the cognitive processing of middle-aged and older adults who may have more life experience and refined self-reflection skills.

Due to the cross-sectional nature of this study, the results should be interpreted as associations, not causal inferences. While the theoretical framework suggests directionality between self-efficacy and self-efficacy sources, this cannot be established through the study design used. Moreover, despite a substantial proportion of the variance explained, 44% remained unexplained, suggesting the presence of unknown and potentially confounding variables. Although sources were measured via a validated questionnaire, the instrument did not contain items assessing physiological states or constructs theorised to moderate levels of self-efficacy derived from sources, such as attribution, task difficulty, source credibility, and model characteristics.

The reliance on self-report measures introduces the potential for recall and social desirability bias, and demand characteristics. Linear regression analysis was employed in the current study, whereas sources and self-efficacy may have curvilinear relationships.^4^ Future studies could address this by incorporating quadratic regression to explore these complexities.

Moreover, in the qualitative inquiry, each question specifically probed a distinct source of self-efficacy, corresponding to those measured within the quantitative questionnaire. This approach likely curtailed theme development and the opportunity to elicit additional potential efficacy sources. In contrast to other qualitative data collection approaches, such as interviews, the use of a structured questionnaire did not facilitate the enrichment of data through the probing of responses.

Despite these limitations, this study had several strengths. It is the first within the PA domain to quantitatively investigate the associations between sources of self-efficacy and self-efficacy among younger adults, compare interaction effects between mastery and other sources, and complement findings with qualitative methods. As such, this study provides evidence on how the sources of self-efficacy are associated with self-efficacy for PA, which could help inform future interventions to reduce inactivity levels. The use of mixed-methods facilitated data triangulation and the elucidation of findings in richer detail than quantitative methods alone.^19^ Another strength was the employment of intercoder agreement using recommended methods,^30^ which fostered improved coding standards, reflexivity, and dialogue within the research team. This process ensured that the coding framework extended beyond the perspective of a single researcher.^30^

Future Directions: Future research should incorporate and test additional potential sources and moderators of sources of self-efficacy. Consistent with Self-Efficacy Theory, these may include physiological states, attributions, source credibility, and model characteristics. In the present study, self-persuasion, a source not originally specified within the tenets of Self-Efficacy Theory, was positively associated with self-efficacy, thereby demonstrating the value of adding and testing additional sources in future research. For instance, relation-inferred self-efficacy,^5^ which concerns an individual’s perceptions of others’ confidence in their abilities, self-modelling through mental imagery, and instructional or negative forms of self-talk, could be incorporated as variables. Moreover, novel sources of self-efficacy may be identified through inductive qualitative approaches, such as grounded theory, and subsequently quantitively tested to ascertain their predictive power. Moreover, the current study’s exploration of interactions was confined to mastery experience and other sources. Future research with larger sample sizes could explore the interactions among more sources of self-efficacy. Additionally, the current findings suggest that for certain individuals, vicarious experience augmented self-efficacy, but undermine it for others. Thus, future research may wish to explore the personal or situational factors that moderate self-efficacy derived from this source. Moreover, experimentally manipulating these sources, either individually or in combination, using a factorial design and manipulation checks, could offer valuable insights into their effects on self-efficacy. Such an approach, characterised by high internal validity, would address some of the methodological limitations of the current study, establish causation, and provide further evidence regarding which sources are most efficacious, and in what combinations, to augment self-efficacy for PA.

## Conclusion

The current study identified mastery experience, self-persuasion, and vicarious experience as being significantly associated with self-efficacy for PA among younger adults. The findings further indicate that self-persuasion and vicarious experience are particularly salient for individuals with lower, compared to higher mastery experience. These findings were generally corroborated by the qualitative data, which also suggested that vicarious experiences can either augment or decrease self-efficacy for PA. To promote self-efficacy, practitioners or researchers should employ BCTs aligned to these identified sources.

## Data Availability

All quantitative and qualitative data files are available upon request to the authors

